# Breakfast skipping as an indicator of unhealthy lifestyle among adolescents: A short letter

**DOI:** 10.1101/2023.04.10.23288359

**Authors:** Mojtaba Daneshvar

## Abstract

Regular breakfast consumption plays a crucial role in the overall health and well-being of children and adolescents, making it an essential factor in maintaining healthy lifestyle habits. This study examined the association between breakfast skipping (BS) and unhealthy lifestyle behaviors in US adolescents using the Youth Risk Behavior Surveillance System (YRBS) 2018 survey. The prevalence of BS was significantly linked to age and race but not gender. After adjusting for confounding variables, a significant positive correlation was observed between BS and low physical activity, smoking, alcohol consumption, and high TV time. No such association was found with high video-game time. The findings highlight the importance of regular breakfast consumption as a vital component of healthy lifestyles among youth. Encouraging regular breakfast habits may reduce the risk of developing unhealthy behaviors and obesity in this population

## Introduction

Regular breakfast eating plays a key role in general health, especially among children and adolescents. The prevalence of breakfast skipping (BS) is reported to be a growing trend in high-income countries [1]. Several studies have underlined the association of BS to the childhood obesity and metabolic disorders. The variety of conditions also extends to psychological alterations and unhealthy lifestyle patterns. It has been demonstrated that breakfast skippers consume regimens with lower nutritional quality, compared to regular breakfast eaters. It has been considered that irregular breakfast consumption is also related to lower physical activity (PA), higher screen time, and substance use. This relationship might be mediated by hormonal and circadian dysregulation in response to BS [1, 2]. With respect to the importance of the BS as a predictor of healthy lifestyle habits, the linkage is not well studied [3].

In this regard, the current study is conducted to evaluate the association between BS with lifestyle risk factors and obesity among US adolescents.

## Method

This data was obtained from the “youth risk behavior surveillance system (YRBS) 2018” survey (https://www.cdc.gov/healthyyouth/data/yrbs/data.htm). YRBS is a cross-sectional national survey which targeted US school students in grades 9-12^th^. A random cluster sampling method was performed and students were asked to participate. YRBS 2019, a questionnaire consisting of 99 questions was utilized to obtain demographic, anthropometrics, lifestyle, behavioral, and school health-related information. Data were obtained using pre-defined response options and recorded as categories (ordinal or nominal). Anthropometric measurements were evaluated based on standard methods, and the Body mass index (BMI) percentile was reported.

Included lifestyle and anthropometric measures and related definitions are listed below:

- BS: eating breakfast during the past 7 days (< 1 day)
- Low PA: having physical activity with a duration of 60 minutes per day (< 5 days)
- High video/computer game time: hours per day (< 3 days)
- High TV time: hours per day (< 3 days)
- Smoking: smoking cigarettes during the past 30 days (>= 1 day)
- Alcohol: having at least one drink of alcohol during the past 30 days (>= 1 day)
- Overweight/obesity: BMI percentile >= 85 (according to BMI-for-Age)

Additional methodological details are available and published previously [4].

All statistical analyses were applied using SPSS 23.0 (SPSS Inc, Chicago, IL, USA), and P-value < 0.05 was considered as significant. Crude and multivariate logistic regression (adjusted for age, sex, and race), were applied to investigate the odds of unhealthy lifestyle factors according to BS categories. Results from logistic regression were expressed as odds ratio (OR) and 95% confidence interval (CI), by considering regular breakfast consumption (>=1 day/week) as reference group.

## Results and discussion

As illustrated in **Table 1**, the prevalence of BS was significantly associated with age group and race (P < 0.001). The prevalence of BS was not different between genders (P = 0.72). Our main analysis was conducted by considering BS as independent factor and (each) unhealthy lifestyle habit as dependent variable (**Table 2**). In the crude model, higher odds of low PA (OR = 1.77), smoking (OR = 1.79), alcohol drinking (OR = 1.28) (P < 0.001), and high TV time (OR = 1.03, P = 0.02) were found among BS groups compared to non-BS group. After adjustment for possible confounding variables (age, sex, and race), the association between BS and these variables remained significant. Higher video-game time was not related to BS in crude, nor adjusted models (P > 0.05).

**Table 1.** General characteristics of study participants across groups of breakfast skipping

| Variables |  | Breakfast skipping |  | P-value |
| --- | --- | --- | --- | --- |
|  |  | Yes | No |  |
| N |  | 1956 (16.9%) | 9637 (83.1%) |  |
| Age group | =< 14 years | 217 (11.2%) | 128 (13.1%) | < 0.001 |
|  | 15, 16 years | 956 (49.2%) | 5036 (52.5%) |  |
|  | >= 17 years | 770 (39.6%) | 3296 (34.4%) |  |
| Sex | Female | 984 (50.9%) | 4904 (51.3%) | 0.72 |
|  | Male | 950 (49.1%) | 4651 (48.7%) |  |
| Race | White | 848 (44.7%) | 4756 (50.8%) | < 0.001 |
|  | African American | 332 (17.5%) | 1260 (13.4%) |  |
|  | Hispanic/ Latino | 167 (8.8%) | 782 (8.3%) |  |
|  | Others | 550 (29.0%) | 2573 (27.5%) |  |

Note: missing data were not reported for categories

**Table 2.**
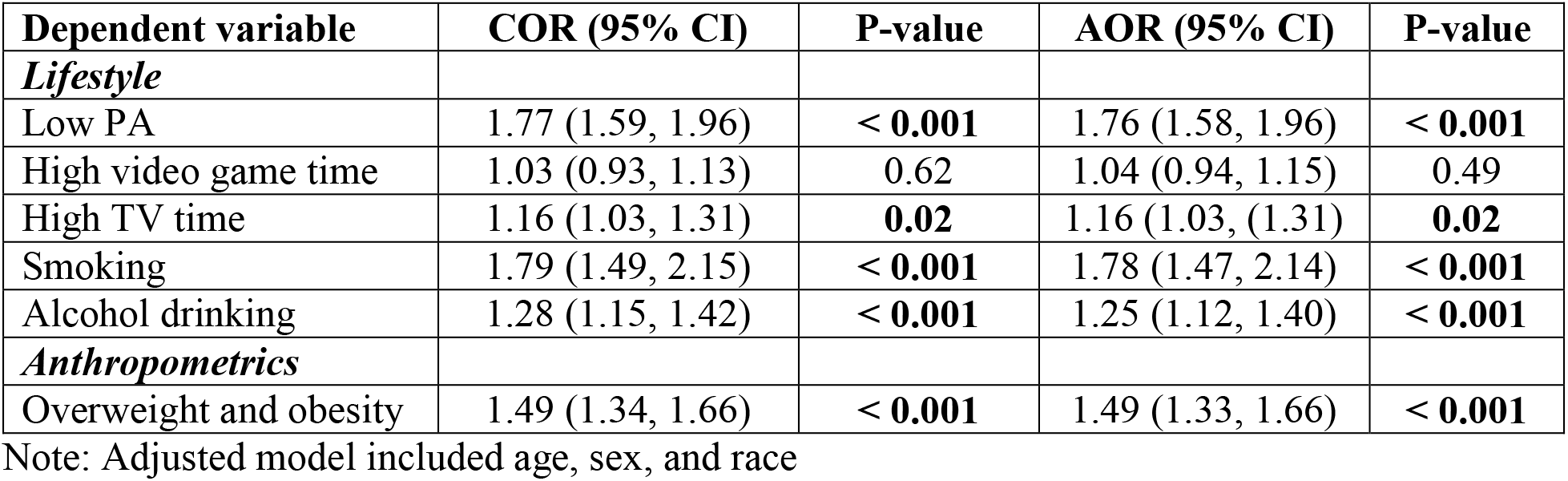
Association between BS and odds of unhealthy lifestyle habits and obesity

Current results provide a positive correlation between BS and unhealthy lifestyle factors (except for high video-game time). This relationship, highlights the importance of regular breakfast consumption among youth; However, the causal association is not established due to the study design. Previous studies have proposed regular breakfast eating as an important meal and an indicator of healthy lifestyle [3, 5]. At this point, our results might support their major assumption and in parallel with this hypothesis. It would be noteworthy to point that the current study aimed to inspect the relationship between lifestyle variables which are conventionally considered as exposure/predictors for major outcomes (metabolic disorders etc.) [5]. Although, results showed a significant association between BS and overweight/obesity, evaluation of this relationship for other metabolic outcomes is out of the scope of this study. In summary, we found that BS could be represented as a substantial element of the lifestyle network. Further studies are needed to support these findings.

## Data Availability

All data produced are available online at:
https://www.cdc.gov/healthyyouth/data/yrbs/data.htm

## Funding

None

## Author Contributions

Designation of the study, statistical analyses, and writing manuscript were performed by M.D.

## Conflict of Interest

None

## Notes

### Competing Interest Statement

The authors have declared no competing interest.

### Author Declarations

Data were openly available before the initiation of the study (https://www.cdc.gov/healthyyouth/data/yrbs/data.htm)

